# Classifying Obsessive-Compulsive Disorder from Resting-State EEG using Convolutional Neural Networks: A Pilot Study

**DOI:** 10.1101/2025.05.06.25327094

**Authors:** Brian A. Zaboski, Sarah Kathryn Fineberg, Patrick D. Skosnik, Stephen Kichuk, Madison Fitzpatrick, Christopher Pittenger

## Abstract

**Objective:** Identifying obsessive-compulsive disorder (OCD) using brain data remains challenging. Resting-state electroencephalography (EEG) offers an affordable and noninvasive approach, but identifying predictive signals in EEG data has met with little success, even with the application of traditional machine learning methods. We explored whether convolutional neural networks (CNNs) applied to EEG time-frequency representations can distinguish individuals with OCD from healthy controls. **Method:** We collected resting-state EEG data from 20 unmedicated participants (10 with OCD, 10 healthy controls). Four-second EEG segments were transformed into time-frequency representations. We then trained a 2D CNN using a leave-one-subject-out cross-validation framework to perform subject-level classification and compared its performance to a more traditional support vector machine (SVM) approach. Next, using multimodal fusion, we examined whether adding clinical and demographic information improved classification. **Results:** The CNN classifier achieved high subject-level performance, distinguishing individuals with an accuracy of 85.0% and an area under the curve (AUC) of 0.88. This significantly outperformed the SVM baseline, which performed no better than chance (45.0% accuracy, AUC: 0.47). A subsequent multimodal analysis revealed that clinical and demographic variables did not contribute any additional independent information. **Conclusion:** CNNs applied to resting-state EEG show promise for identifying OCD, outperforming traditional machine learning methods. These findings highlight the potential of deep learning to uncover complex, diagnostically relevant patterns in neural data. While limited by sample size, this work supports further investigation into multimodal models for psychiatric classification, warranting replication in larger, more diverse samples.

---

Obsessive-Compulsive Disorder (OCD) is a prevalent and often debilitating neuropsychiatric condition, affecting 1-3% of the population worldwide and causing significant functional impairment and distress (Kessler et al., 2012; Ruscio et al., 2010). While effective treatments exist, including cognitive-behavioral therapy (CBT) and pharmacotherapy with selective serotonin reuptake inhibitors (SSRIs), a substantial portion of patients do not achieve remission (Abramowitz et al., 2010; Law & Boisseau, 2019). Diagnosis currently relies on clinical interviews and standardized symptom scales, such as the Yale-Brown Obsessive-Compulsive Scale, which capture subjective reports of obsessions and compulsions (Goodman et al., 1989). The inherent subjectivity in this process, alongside the complexities in differentiating certain symptom presentations (Mattera et al., 2024), highlights a critical need for new models to aid in diagnosis and predict treatment response (Zaboski et al., 2021).

Neuroimaging modalities like positron emission tomography (PET) and functional magnetic resonance imaging (fMRI) have identified new treatment approaches (Rance et al., 2023) as well as functional and structural alterations in individuals with OCD, particularly within cortico-striatal-thalamo-cortical circuits (Pittenger, 2017; Stein et al., 2019). However, the high cost, limited accessibility, and relatively poor temporal resolution of these techniques hinder their widespread clinical application (Constable, 2023). Moreover, mega-analyses using whole-brain functional connectivity have struggled to find strong individual classification performance (Bruin et al., 2023). Electroencephalography (EEG) offers a non-invasive, relatively inexpensive, portable method for measuring the brain’s electrical activity, with millisecond temporal resolution (Zaboski et al., 2021). Despite these advantages, the use of EEG to investigate OCD has been comparatively sparse, with studies yielding heterogeneous findings regarding event-related potentials (event-related potentials [ERPs] like the P300 and the error-related negativity [ERN]), oscillatory power abnormalities (e.g., in theta, alpha, and beta bands), and measures of signal complexity (Metin et al., 2019; Zaboski et al., 2021).

The complexity and high dimensionality of EEG data present challenges for traditional analytic methods. Machine learning (ML) approaches have emerged as powerful tools for identifying subtle patterns within complex biological data. Previous studies in OCD have employed ML techniques, often using support vector machines (SVMs) or conventional artificial neural networks (ANNs). A common characteristic of these approaches was their application to pre-defined quantitative EEG (qEEG) features—like spectral band power or cordance—that were engineered from the EEG signals prior to model training. For example, Metin et al., (2019) found that theta band power derived from qEEG (analyzed with ANNs) could predict TMS response. Similarly, Erguzel et al. (2015) used SVMs on qEEG cordance to classify OCD versus trichotillomania. While these methods have shown utility, their dependence on manual feature engineering is both cumbersome and may not fully capture the intricate patterns embedded in EEG data, a limitation that modern deep learning addresses.

More recently, convolutional neural networks (CNNs) have shown remarkable success in automatically extracting informative hierarchical features from raw or minimally processed data, bypassing manual feature engineering (Farhad et al., 2024; Xu et al., 2018; Zaboski et al., 2025). CNNs are well suited for analyzing grid-like data structures, such as the time-frequency representations of EEG signals or even the raw multichannel time-series data itself (Farhad et al., 2024; Xu et al., 2018). Consequently, their application in EEG is growing rapidly for various neurological and psychiatric conditions (Farhad et al., 2024). However, few studies—and none in OCD—have applied deep learning to automatically learn discriminative patterns directly from high-dimensional resting-state EEG data (Zaboski et al., 2025).

To address this gap, we investigated the feasibility of using a CNN to capture the richness of resting state EEG’s temporal signals to classify individuals diagnosed with OCD from healthy controls. By applying CNNs to resting state signals, we hope to bypass manual feature engineering (such as calculating power in specific bands) while capturing complex spatio-temporal and spectral dynamics missed by simpler models or feature sets. We tested whether a CNN can outperform a more conventional machine learning approach to classification. We then systematically added clinical and demographic predictors to the CNN model to determine if its predictive metrics would improve. Success in this endeavor could help us develop more objective, scalable, and accessible tools for OCD research and clinical practice, aligning with the broader goal of achieving precision medicine in psychiatry (Zaboski & Bednarek, 2025).

## Methods

### Participants

Participants were recruited from the local community for a larger 18-week clinical trial investigating brain network changes and treatment prediction in response to pharmacotherapy for OCD (“Brain network changes accompanying and predicting treatment responses to pharmacotherapy in OCD”, Yale HIC#: 2000023688; PI: C. Pittenger). Recruitment methods included social media advertisements, local bus ads, community flyers, and referrals to the Yale OCD Research Clinic. The study protocol was approved by the Yale University Institutional Review Board (IRB), and all participants provided written informed consent prior to participation, including specific consent for the EEG procedures described herein. Participants received $50 for completing the baseline clinical/self-report assessment battery and an additional $80 for the baseline EEG session.

Participants underwent an initial screening followed by a comprehensive intake evaluation (in-person or remote). The primary diagnosis of OCD was established using either the Mini-International Neuropsychiatric Interview (MINI; Sheehan et al., 1998) or Diagnostic Interview for Anxiety, Mood, and OCD and Related Neuropsychiatric Disorders (DIAMOND; (Tolin et al., 2018) and validated by a board-certified psychologist or psychiatrist associated with the study. The same diagnostic process was used for healthy control (HC) participants. HCs were excluded if they had any current DSM-5 diagnosis, including symptoms of psychosis, mania, depression, or substance use.

We performed medical screening for OCD participants as part of the parent study, which included assessment of standard laboratory blood tests (including complete blood count, metabolic panel, thyroid tests), urine tests (for kidney function and drug screen), electrocardiogram (ECG), and a physical examination to ensure general health. Key exclusion criteria relevant to the larger study included pregnancy or breastfeeding, magnetic resonance imaging (MRI) contraindications (e.g., certain metallic implants), current use of specific interacting medications (e.g., monoamine oxidase inhibitors, certain serotonergic agents, warfarin unless specifically approved), and significant suicide risk as determined by the investigators. Participants agreed not to start new medications during the study period.

Baseline EEG data were analyzed for the present study. Participants completed self-report scales assessing mood, OCD symptoms, personality characteristics, and other relevant factors using REDCap, a secure, web-based, HIPAA-compliant data-capture system (P. A. Harris et al., 2009, 2019) and a baseline EEG session. 22 participants (11 OCD, 11 HC) completed the baseline EEG session. Initial preprocessing yielded unusable data for two participants (one HC, one OCD). Despite artifact correction attempts including ICA two individuals exhibited high-amplitude noise surpassing the 250 µV rejection criterion, necessitating their exclusion from the final analysis. *N* = 20 participants (*n* = 10 OCD, *n* = 10 HC) were retained for the final analysis.

### Data Acquisition

Resting-state EEG data were acquired within the Psychophysiology Laboratory in the Connecticut Mental Health Center. The laboratory is specifically designed for acquiring high-quality human psychophysiology data and features an electromagnetically and acoustically shielded testing booth (8 x 6 x 9 ft) to minimize environmental noise and electrical interference during recording. Within these booths, participants were seated comfortably in a reclining chair.

The EEG recording system utilized was a 64-channel Biosemi ActiveTwo system. Data were collected using a standard 64-channel electrode cap, arranged according to the international 10-20 system. The recordings used an average reference; additional bipolar electrode pairs were employed to record vertical and horizontal electrooculogram (EOG) signals to monitor eye movements and blinks for subsequent artifact processing. Throughout the recording sessions, electrode impedances were maintained below 10 kΩ to ensure good signal quality. The EEG data were continuously sampled at a rate of 1024 Hz and acquired with an online hardware band-pass filter set at 0.1–100 Hz. Recordings were made under eyes open conditions for 4 minutes at a sampling rate of and saved in .bdf format.

### Data Preprocessing

We preprocessed the raw EEG data using the MNE-Python library (v1.7) (Gramfort, 2013; Gramfort et al., 2014) to remove artifacts and prepare the signals for analysis. Initially, raw data files were loaded, and any non-EEG or status channels were removed. The standard 10-20 montage configuration was applied. We applied a 60 Hz notch filter to remove power line interference, followed by a band-pass filter between 0.5 Hz and 50.0 Hz (FIR filter) to eliminate slow drifts and high-frequency noise. Each participant’s recording was then cropped to a uniform duration of 240 seconds to standardize the data length across the sample.

Artifacts primarily related to eye movements and blinks were addressed using independent component analysis (ICA). We fitted the FastICA algorithm to the filtered data (Aapo, 1999; Ablin et al., 2018; Bell & Sejnowski, 1995; Lee et al., 1999) to derive 15 independent components. Components exhibiting high correlation with simultaneously recorded EOG channels were automatically identified and removed. On average, 0 – 2 were removed per participant. No specific procedure for removing ECG-related artifacts was implemented; however, our subsequent epoch rejection step would have removed segments with significant cardiac artifacts. The EEG signal was then reconstructed from the remaining non-artifactual components. Reconstructed EEG data were re-referenced to the common average across all channels (Jiang et al., 2024; Tamburro et al., 2021). Finally, the cleaned, continuous data were segmented into 4-second epochs with a 2-second overlap between consecutive epochs. This initial segmentation yielded 190 epochs per subject, for a total of 3,800 epochs across the 20 participants. To remove any remaining high-amplitude artifacts not corrected by ICA, an automatic rejection procedure was applied. Any epoch where the peak-to-peak signal amplitude of any EEG channel exceeded 300 µV was discarded. This automatic rejection procedure resulted in a mean of 113.8 ± 5.9 clean epochs per participant (range: 101 to 119). We performed the subsequent time-frequency transformation on the remaining 2,277 clean epochs.

### Time-Frequency Feature Generation

To capture dynamic neural activity patterns suitable for the CNN, the preprocessed EEG epochs were transformed into time-frequency representations. This was achieved using a Morlet wavelet transform implemented in MNE-Python, applied independently to each EEG channel within each epoch. Time-frequency representations were computed for each 4-second epoch using MNE-Python’s tfr_morlet function. To preserve the original signal and prevent data loss, no baseline correction was applied (Urbach & Kutas, 2006). We analyzed 40 logarithmically spaced frequencies ranging from 1 Hz to 45 Hz. The number of cycles for each wavelet was set proportionally to the frequency (n_cycles = frequency / 2.0). To manage computational load and data size, the resulting TFR output was decimated by a factor of 16. Given the original 1024 Hz sampling rate resulted in 4096 samples per 4-second epoch, this decimation yielded a final temporal dimension of 256 time points per epoch for CNN input. The resulting absolute power values were then log-transformed (base 10, scaled by 10, adding 1e-10 for stability) to approximate a normal distribution and reduce the influence of extreme values. The subsequent time-frequency transformation resulted in one multi-channel power ‘image’ per epoch, totaling 2,277 images. Each image had the dimensions 40 frequencies, 256 time points, 68 channels. These images served as the input features for CNN classification.

### Primary Classification Procedure

Our primary goal was to classify individual subjects. To achieve this with a convolutional neural network despite the limited sample size, we adopted a two-stage approach involving epoch-level training followed by subject-level evaluation.

### Epoch-Level Training Strategy and Rationale

First, to provide a CNN with sufficient training examples, we trained it to classify individual EEG epochs. We used it to classify individual EEG epochs as belonging to either the HC or OCD group. This strategy leverages the multiple epochs from each participant (∼114 each) as a form of data augmentation, creating a large set of training instances from the limited subject pool. We acknowledge that epochs from the same subject are not statistically independent, a common challenge in this type of analysis. To rigorously control for individual subject effects and to prevent the model from gaining access to data outside the training set, we applied leave-one-subject-out (LOSO) cross-validation (Mevlevioğlu et al., 2024; Ren et al., 2024). Across the 20 folds of this procedure, the model was trained on data from 19 subjects and evaluated on all epochs from the held-out subject. Data scaling (StandardScaler) was applied within each fold, fitted only on the training data.

### CNN Architecture

The classification model utilized a two-dimensional CNN implemented using the Keras API (Chollet & others, 2015) within TensorFlow (Martín Abadi et al., 2015). The architecture consisted of two sequential convolutional blocks: The first block employed 16 filters with a (3, 5) kernel size, a choice theoretically motivated to capture local patterns across adjacent frequency bins and short temporal dynamics, followed by ReLU activation, batch normalization, (2, 2) max-pooling, and dropout (rate = 0.4). The second block used 32 filters with a (3, 5) kernel size and identical subsequent layers (ReLU, batch normalization, max-pooling, dropout). Following the convolutional blocks, the feature maps were flattened and passed directly to a single dense output neuron with a sigmoid activation function. This final layer produces a continuous output between 0 and 1 for each epoch, representing the model’s predicted probability of that epoch belonging to the OCD class. The architecture incorporated several regularization techniques (batch normalization, dropout) to mitigate the risk of overfitting, a key consideration given the study’s sample size. The computed log-power time-frequency representations for each epoch were reshaped to conform to the ‘channels-last’ input format expected by the 2D CNN. Specifically, the dimensions were permuted from (epochs, channels, frequencies, time points) to (epochs, frequencies, time points, channels). The resulting input tensor had dimensions 40 (Frequencies) × 256 (Time Points) × 68 (EEG Channels). The model was trained using the Adam optimizer with a learning rate of 0.001 and a batch size of 32, which are common and robust default values for models of this type (Kingma & Ba, 2017; Krizhevsky et al., 2012). Average training and validation loss curves across the 20 folds are provided in the Supplement, demonstrating successful model convergence.

### Subject-Level Classification

The direct output of the CNN’s final sigmoid layer was a probability for each individual epoch. To derive a single, robust prediction for each participant, we aggregated these epoch-level probabilities. For each held-out subject in the LOSO procedure, we calculated the mean of the predicted probabilities across all of their test epochs. This average probability served as the final subject-level EEG score. A subject was classified as having OCD if their score was greater than 0.5 and as a HC otherwise. The model’s primary performance metrics, including accuracy, AUC, and the confusion matrix, were then calculated based on these 20 subject-level predictions.

### Support Vector Machine Baseline Comparison

To contextualize the performance of the CNN model, we conducted a baseline comparison using a standard support vector machine (SVM) with traditional EEG features (Bera et al., 2025; Rahul et al., 2024) also predicting group epochs. The SVM classifier with a radial basis function (RBF) kernel (C = 1.0, gamma = ‘scale’, class_weight = ‘balanced’) was trained on features representing the mean spectral power within canonical frequency bands (delta: 1 – 4 Hz, theta: 4 – 8 Hz, alpha: 8 – 13 Hz, beta: 13 – 30 Hz, gamma: 30 – 45 Hz) calculated for each channel using Welch’s method. To ensure direct comparability, an identical LOSO cross-validation procedure, including per-fold data scaling, was employed.

### Input Visualization

To provide a concrete, illustrative example of the time-frequency features available to the CNN, Figure 1 contains the average power representation for the Cz electrode from a representative HC and OCD participant. We selected the Cz electrode for this visualization because, as a central midline site, it often reflects widespread neural dynamics and is a common choice in EEG research for providing a stable and representative view of overall spectral activity.

**Figure 1:**
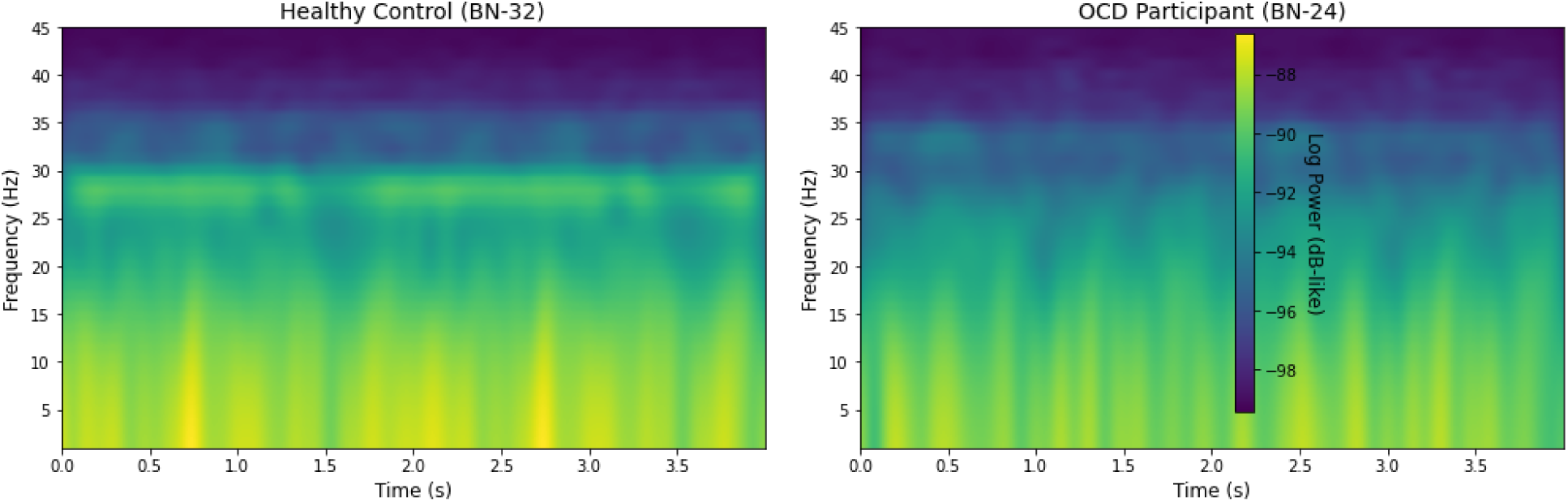
Example Average Time-Frequency Power (Cz) for Two Subjects.

Visual inspection of these representative subjects revealed distinct patterns. The HC participant (BN-32) exhibited prominent, sustained spectral power predominantly within the high-beta frequency band (∼20-30 Hz). In contrast, the OCD participant (BN-24) displayed lower overall power and notably lacked this sustained high-beta activity.

Figure 1 highlights the theoretical advantage of the CNN approach. While the figure illustrates clear differences in the dynamics of beta activity, a traditional machine learning model like an SVM trained on mean spectral power would reduce this rich pattern to a single number per frequency band, losing all temporal information (Chaddad et al., 2023; Hosseini et al., 2020; Lotte et al., 2007; Salehzadeh et al., 2023). It is crucial to recognize that our CNN did not operate on these averaged, single-channel representations (Schirrmeister et al., 2017). Instead, it processed the complete, multi-channel data for each individual 4-second epoch.

The structure of the CNN’s input data is key. Analogous to a standard color image, which is a single entity of [height, width, 3 color channels] (Krohn et al., 2019), the data for a single epoch fed into our CNN was a single entity of [40 Frequencies (Height), 256 Time Points (Width), 68 EEG Channels (Depth)]. By analyzing this multi-channel entity, the network could learn patterns based not only on the time-frequency content within a single channel but also on the spatial relationships between all 68 channels across the scalp (Zhang et al., 2023).

### Multimodal Classification Procedure

To determine if clinical and demographic information provided additional diagnostic value, we conducted a second-stage analysis investigating the addition of clinical and demographic information to the CNN-derived EEG score using a late-fusion (stacking) approach (Breiman, 1996; Wolpert, 1992). In this stage, we predicted subject status. Output of the

second-stage represented a probability, indicating confidence in the subject’s classification. Similar to Stage One, the final binary prediction (0 or 1) was derived by thresholding this probability at 0.5.

This analysis began by computing subject-level EEG scores, obtained by averaging the predictions on the held-out subject from each fold of the primary CNN’s LOSO evaluation. Next, subject-level EEG scores were merged with clinical scores (e.g., DOCS, BAI, and BDI-II totals) and demographic data (age, sex, education), matched by participant ID. To systematically assess the contribution of individual variables while minimizing overfitting, we tested the addition of each clinical or demographic feature one at a time to the EEG score (Baltrušaitis et al., 2018).

For each feature combination tested, a Scikit-learn (Pedregosa et al., 2011) pipeline was created to preprocess the two input features within the cross-validation loop: the numeric EEG score (and any other numeric variable being tested, like age) was standardized using StandardScaler. Any categorical variable being tested (e.g., sex, education) was one-hot encoded. A logistic regression classifier was selected as the second-stage model, and each combined model (feature pair + preprocessing + logistic regression) was evaluated using the identical LOSO cross-validation procedure employed for the original CNN. This ensured equitable comparison across feature combinations.

Lastly, in addition to using subject-level scores from the CNN, we also generated scores from the support vector machine. We then tested each variable as described above to test if the SVM prediction would improve. If the SVM led to significantly greater performance, this would imply that there is information in the clinical and demographic features that is already captured in the EEG signal as analyzed using the CNN.

### Statistical Analysis and Software

The primary performance metrics for evaluating both the EEG-only CNN and the combined multimodal model were the area under the receiver operating characteristic curve (AUC) and overall classification accuracy (Naidu et al., 2023). Performance was assessed both per fold and globally by aggregating predictions across all LOSO folds. Overall confusion matrices and classification reports detailing precision, recall, and F1-scores were generated. Average performance across folds is reported as mean ± standard deviation. The analysis used Python (Van Rossum & Drake, 2009) and key scientific libraries including MNE-Python (v1.7) (Gramfort, 2013; Gramfort et al., 2014), NumPy (C. R. Harris et al., 2020), Pandas (McKinney, 2010), Scikit-learn (Pedregosa et al., 2011), TensorFlow (Martín Abadi et al., 2015) with Keras (Chollet & others, 2015), Matplotlib (Hunter, 2007), and Seaborn (Waskom, 2021).

### Clinical Measures

#### Dimensional Obsessive-Compulsive Scale

The Dimensional Obsessive-Compulsive Scale (DOCS) is a 20-item self-report tool created to assess the severity of Obsessive-Compulsive Disorder (OCD) symptoms across four well-established dimensions: contamination concerns paired with cleaning compulsions, fears of responsibility for harm leading to checking behaviors, unacceptable thoughts accompanied by mental rituals, and symmetry obsessions linked with ordering compulsions (Abramowitz et al., 2010). Initial psychometric evaluations indicated that the DOCS possesses strong internal consistency (α ≈ 0.90). Its capacity for detecting OCD symptoms (diagnostic sensitivity) is further supported by a strong positive correlation (*r* = 0.69) between DOCS total scores and scores on the Obsessive-Compulsive Inventory-Revised (Abramowitz et al., 2010; Abramowitz & Deacon, 2006).

#### Magical Ideation Scale

The Magical Ideation Scale (MIS) is a 30-item self-report instrument developed to measure unlikely causal beliefs, such as telepathy, superstitious thinking, and thought broadcasting (Eckblad & Chapman, 1983). It employs a true/false response format where respondents endorse statements reflecting personal beliefs and experiences related to these phenomena (e.g., “I have sometimes felt that strangers were reading my mind”) (Eckblad & Chapman, 1983). The total score, derived by summing the items keyed towards magical ideation, serves as an indicator of this cognitive style, which is considered a feature of schizotypy or psychosis-proneness (Eckblad & Chapman, 1983; Kwapil et al., 1997). Psychometric studies have generally shown good internal consistency, with reliability coefficients often reported between *r* = .76 and *r* = .93 across various samples, including adolescents and adults (Fonseca Pedrero et al., 2009; Kingdon et al., 2012). The MIS demonstrates construct validity through correlations with other measures of schizotypy, particularly positive symptoms like perceptual aberrations, and its ability to predict psychotic-like experiences or distinguish individuals at higher risk for psychosis-spectrum disorders in longitudinal studies (Chapman et al., 1982; Fonseca Pedrero et al., 2009; Kwapil et al., 1997; Martin et al., 2011).

#### Beck Anxiety Inventory

The Beck Anxiety Inventory (BAI) is a 21-item self-report questionnaire used to evaluate the severity of anxiety symptoms in adults. Total scores range from 0 to 63. Respondents rate how much they have been bothered by common anxiety symptoms over the past week using a 4-point Likert scale (0 = “Not at all” to 3 = “Severely”) (Beck et al., 1988, 2012). The BAI focuses particularly on the somatic or physical symptoms of anxiety, such as sweating, trembling, and dizziness. The BAI has demonstrated strong internal consistency (α = .91) and test-retest reliability (*r* = .65) in meta-analyses of its psychometric properties (Bardhoshi et al., 2016).

#### Beck Depression Inventory-II

The Beck Depression Inventory-II (BDI-II) is a self-report questionnaire consisting of 21 items designed to gauge the intensity of cognitive and physical symptoms commonly associated with depression (Beck et al., 1961). Respondents typically choose from four statements per item, each reflecting a different level of symptom severity and carrying a specific point value. The sum of these points yields a total score, falling between 0 and 63. This total score is used to classify depression severity into categories: minimal (0 – 13), mild (14 – 19), moderate (20 – 28), or severe (29 – 63). Studies support the BDI-II’s psychometric properties, demonstrating high internal consistency (α ≈ 0.90), adequate test-retest reliability (α ≈ 0.73 – 0.96), and effectiveness in distinguishing between psychiatric patients diagnosed with depression and those without (Beck et al., 1996; Wang & Gorenstein, 2013).

## Results

### Descriptive Statistics

Descriptive statistics on our 20 participants (10 HC and 10 OCD) are presented in Table 1. The average participant age was in the mid-thirties (*M* = 35.4, *SD* = 13.5), with similar distributions between the HC and OCD groups. Regarding clinical scores, mean symptom severity varied across measures: DOCS and BDI-II scores were higher in the OCD group, while MIS scores were slightly lower. BAI scores were comparable between groups.

**Table 1.** Descriptive Statistics for Demographics and Clinical Characteristics.

| Continuous Variable | Overall ($M$, $SD$) | OCD ($M$, $SD$) | HC ($M$, $SD$) |
| --- | --- | --- | --- |
| Age, years | 35.42 (13.46) | 34.89 (14.41) | 35.90 (13.31) |
| DOCS Total | 17.00 (16.98) | 20.80 (17.99) | 13.20 (15.91) |
| MIS Score | 7.00 (6.33) | 5.90 (5.92) | 8.38 (6.97) |

| BAI Total | 13.55 (13.34) | 13.30 (11.87) | 13.80 (15.32) |
| --- | --- | --- | --- |
| BDI-II Total | 11.50 (13.34) | 16.00 (14.50) | 7.00 (10.98) |
| **Categorical Variables** | ***N* (%)** | ***n* (%)** | ***n* (%)** |
| *Sex* |  |  |  |
| Male | 13 (65.0%) | 6 (60.0%) | 7 (70.0%) |
| Female | 7 (35.0%) | 4 (40.0%) | 3 (30.0%) |
| *Education Level* |  |  |  |
| Some College | 2 (10.0%) | 2 (20.0%) | 0 (0.0%) |
| Bachelor Degree | 2 (10.0%) | 2 (20.0%) | 0 (0.0%) |
| Master's Degree | 1 (5.0%) | 0 (0.0%) | 1 (10.0%) |
| Doctoral Degree | 6 (30.0%) | 4 (40.0%) | 2 (20.0%) |
| Other/Professional | 9 (45.0%) | 2 (20.0%) | 7 (70.0%) |

*Note:* $N = 20$ total; $n = 10$ participants in each group. HC = Healthy Control; OCD = Obsessive-Compulsive Disorder; DOCS: Dimensional Obsessive-Compulsive Scale; MIS: Magical Ideation Scale; BAI: Beck Anxiety Inventory; BDI-II: Beck Depression Inventory-II.

The sample consisted predominantly of males (65.0%). A high level of educational attainment characterized the cohort, with 75% of participants holding doctoral degrees or other/professional qualifications; notable differences in the distribution of specific education levels were observed between the HC and OCD groups. Data completeness was high, with missing values limited to age for one participant and MIS scores for two participants.

### Primary Model

#### CNN-Based Subject-Level Classification

The primary evaluation of the CNN was conducted at the subject-level to assess its clinical utility and generalizability to unseen individuals. By averaging the prediction probabilities across all epochs for each held-out participant, we derived a single EEG score per subject for classification that dichotomized an individual as having OCD or not.

This subject-level analysis yielded an overall classification accuracy of 85.0%. The primary metric for discriminative performance, the ROC AUC, was 0.88, indicating strong model performance. The model demonstrated a balanced ability to identify both patient and control groups, with a precision of 0.82 and recall of 0.90 for the OCD group, and a precision of 0.89 and recall of 0.80 for the HC group. Detailed performance metrics are presented in Table 2.

**Table 2.** Subject-Level Classification Performance of the CNN.

| Class | Precision | Recall | F1-Score | Support |
| --- | --- | --- | --- | --- |
| HC (0) | 0.89 | 0.80 | 0.84 | 10 |
| OCD (1) | 0.82 | 0.90 | 0.86 | 10 |
| **Accuracy** | - | - | 0.85 | 20 |
| **Macro Avg** | 0.85 | 0.85 | 0.85 | 20 |
| **Wtd Avg** | 0.85 | 0.85 | 0.85 | 20 |

The model’s subject-level performance is illustrated in Figure 2. The confusion matrix details the specific classification outcomes across all 20 participants: 8 True Negatives (HC correctly identified), 9 True Positives (OCD correctly identified), 1 False Negative (OCD misclassified as HC), and 2 False Positives (HC misclassified as OCD). The ROC curve confirms the model’s ability to distinguish between individuals with OCD and healthy controls.

**Figure 2:**
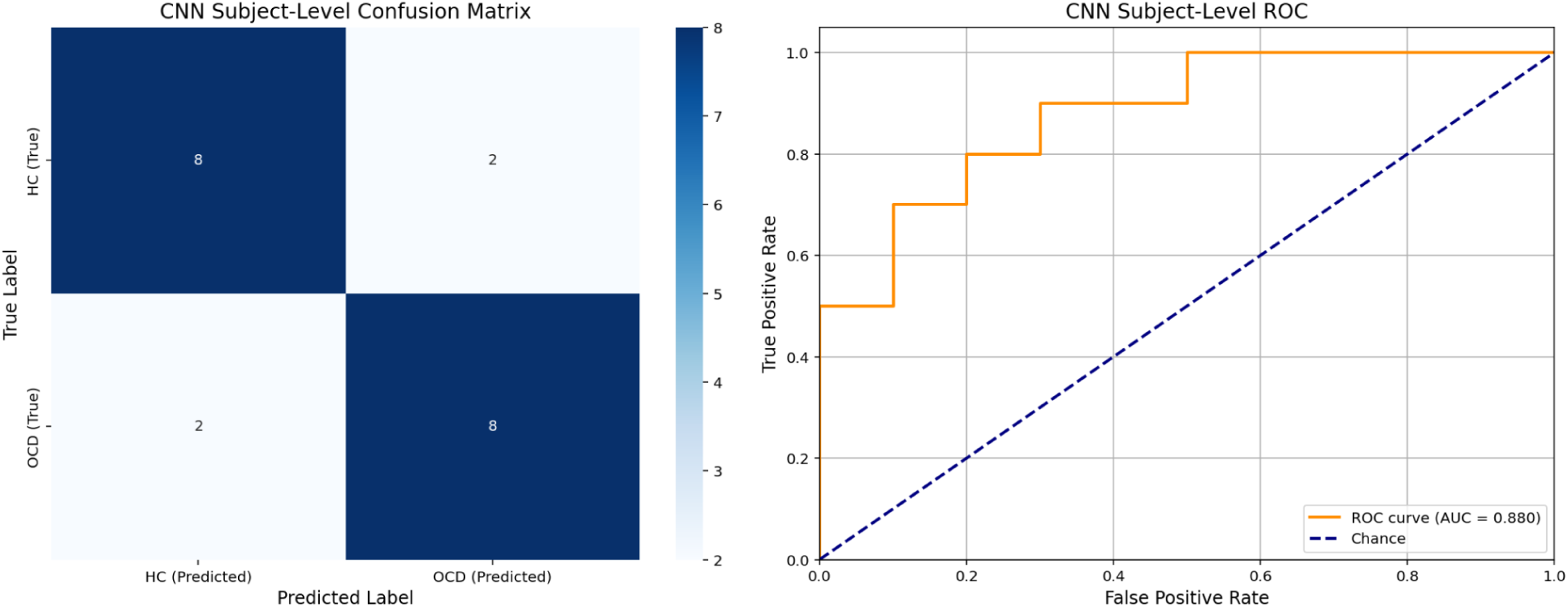
Subject-Level CNN Performance. (Left) The confusion matrix for subject-level predictions (*N* = 20). (Right) The Receiver Operator Curve (ROC) for the subject-level EEG scores, with an Area Under the Curve (AUC) of 0.88.

#### Derivation from Epoch-Level Predictions

We derived these subject-level results from the model’s initial predictions on individual data segments. When aggregated across all 2,277 test epochs, the overall epoch-level accuracy was 81.95% (ROC AUC = 0.86; see Supplementary Table S1 for detailed epoch-level metrics). However, this aggregate metric masked significant performance variability across individuals (mean per-fold accuracy = 81.29% ± 32.17%). This heterogeneity highlights the importance of the subject-level evaluation as the most comprehensive and clinically relevant assessment of the model’s generalizability. These findings strongly suggest that the time-frequency patterns processed by the CNN contain significant, diagnostically relevant information.

#### Support Vector Machine Classification

We constructed an SVM trained on spectral power features (delta: 1 – 4 Hz, theta: 4 – 8 Hz, alpha: 8 – 13 Hz, beta: 13 – 30 Hz, gamma: 30 – 45 Hz). To ensure a fair and direct comparison with our primary CNN analysis, we evaluated the SVM’s performance at the subject level, following the same procedure of averaging epoch-level probabilities to derive a single score per participant. The detailed results of the intermediate epoch-level SVM model, from which these subject-level scores were derived, are provided in the supplement (see supplementary Table S2 and supplementary Figure S3).

This subject-level analysis revealed that the SVM performed no better than chance, achieving an overall accuracy of only 45.0% and an ROC AUC of 0.47 (Table 3; Figure 3). The model’s poor performance, detailed in Table 3 and illustrated in Figure 3, indicates that the traditional band-power features contained limited generalizable information for discriminating between individuals with OCD and healthy controls in this dataset.

**Figure 3:**
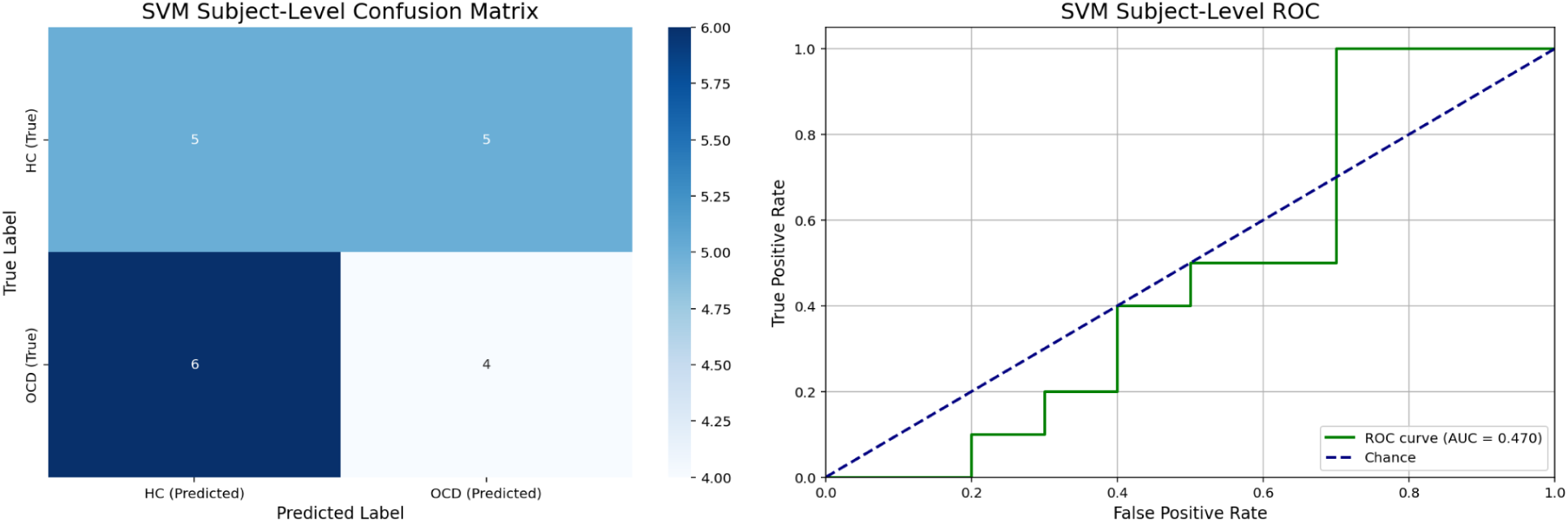
Subject-Level SVM Performance. (Left) The confusion matrix for subject-level predictions (*N* = 20). (Right) The receiver operator curve for the subject-level EEG scores, with an area under the curve (AUC) of 0.47, indicating chance-level performance.

**Table 3.** Subject-Level Classification Performance of the SVM.

| Class | Precision | Recall | F1-Score | Support |
| --- | --- | --- | --- | --- |
| HC (0) | 0.45 | 0.50 | 0.47 | 10 |
| OCD (1) | 0.44 | 0.40 | 0.42 | 10 |
| **Accuracy** | - | - | 0.45 | 20 |
| **Macro Avg** | 0.45 | 0.45 | 0.45 | 20 |
| **Wtd Avg** | 0.45 | 0.45 | 0.45 | 20 |

Notes: Performance metrics are calculated based on one prediction per subject ( $N = 20$ ).

### Multimodal Clinical and Demographic Classification

#### CNN Augmentation

To determine if classification accuracy could be enhanced, we implemented a second-stage analysis using a late-fusion approach incorporating individual clinical and demographic variables. For the multimodal analysis, a baseline model using only the CNN-derived EEG score yielded an AUC of 0.86 and an accuracy of 80.0%. This performance differs slightly from the primary analysis in Table 2, as it reflects the result of a logistic regression model trained on the EEG scores within the cross-validation loop, rather than simple thresholding. The performance metrics for models incorporating clinical scores (DOCS, MIS, BAI, BDI-II) and demographic variables (age, sex, education) are detailed in Table 4.

**Table 4:** Predictive Performance for CNN + Clinical and Demographic Features.

| Feature | *N* | AUC | Accuracy | Precision | Recall | F1 | Change (AUC) |
| --- | --- | --- | --- | --- | --- | --- | --- |
| Baseline EEG | 20 | 0.86 | 0.80 | 0.80 | 0.80 | 0.80 | - |
| DOCS Total | 20 | 0.86 | 0.80 | 0.80 | 0.80 | 0.80 | None |
| MIS Score | 18 | 0.81 | 0.78 | 0.75 | 0.75 | 0.75 | Worse |
| BAI Total | 20 | 0.86 | 0.80 | 0.80 | 0.80 | 0.80 | None |
| BDI-II Total | 20 | 0.84 | 0.80 | 0.80 | 0.80 | 0.80 | Worse |
| Age | 19 | 0.76 | 0.79 | 0.80 | 0.80 | 0.80 | Worse |
| Sex | 20 | 0.81 | 0.80 | 0.80 | 0.80 | 0.80 | Worse |
| Education | 20 | 0.84 | 0.80 | 0.80 | 0.80 | 0.80 | Worse |

*Note:* Baseline metrics include only the convolutional neural network’s subject-level EEG scores in the second stage. *N* = number of subjects remaining after handling missing values. DOCS: Dimensional Obsessive-Compulsive Scale; MIS: Magical Ideation Scale; BAI: Beck Anxiety Inventory; BDI-II: Beck Depression Inventory-II.

The addition of most variables—symptom scores (DOCS, MIS), anxiety/depression scores (BAI, BDI-II), participant age, and sex—did not improve classification performance beyond the baseline EEG CNN model. Several of these combinations resulted in slightly lower AUC values (range 0.76 - 0.86) while maintaining similar overall accuracy (∼80%).

#### SVM Augmentation

Separately, we conducted an analogous multimodal analysis starting from the subject-level score derived from the baseline SVM classifier. This SVM-only baseline, evaluated using the same LOSO framework, performed poorly (AUC = 0.47, Accuracy = 45.0%), indicating limited discriminative information captured from the traditional band power features at the subject level. The results of systematically adding individual clinical and demographic features to the SVM’s baseline EEG score are presented in Table 5.

**Table 5:**
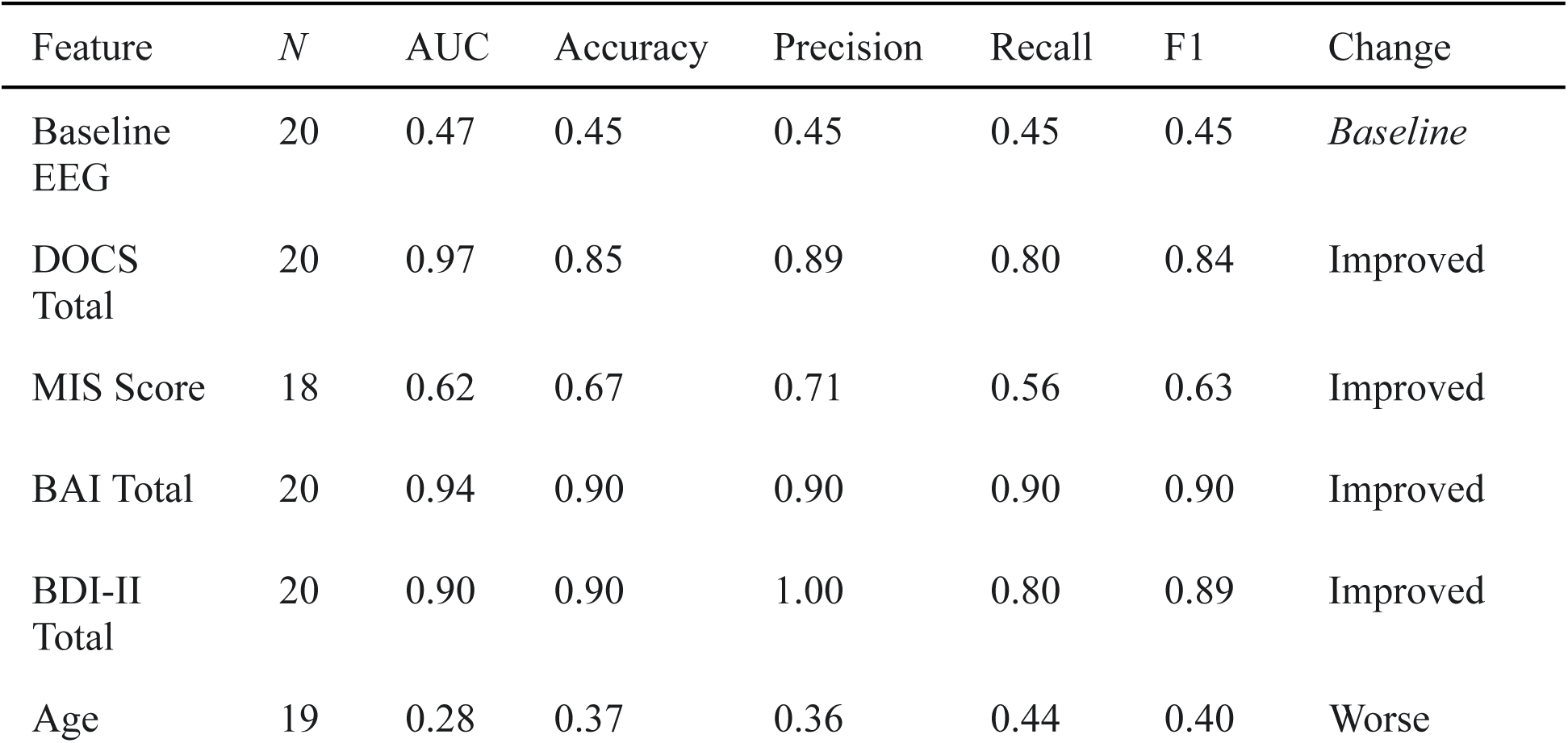
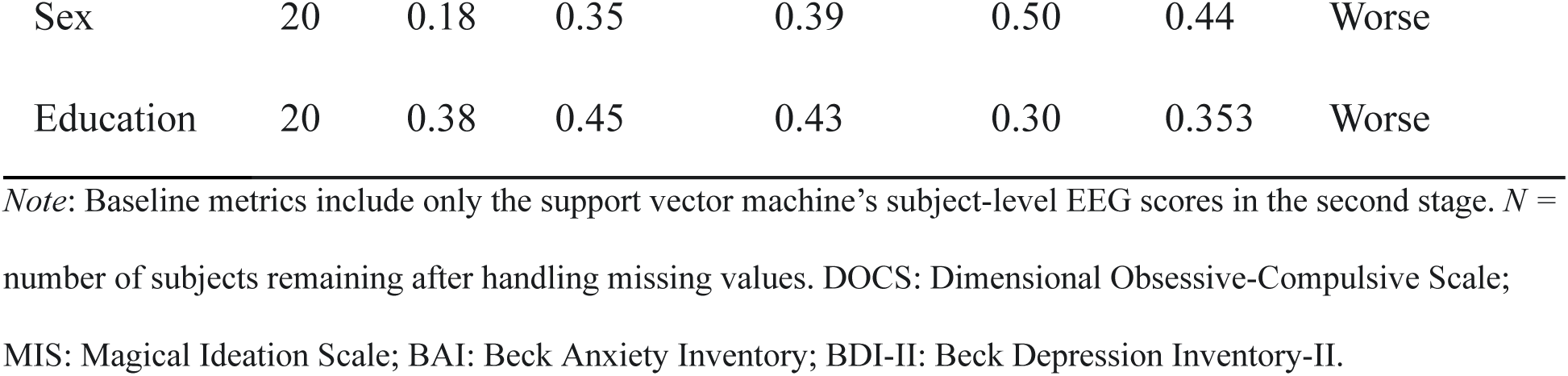
Predictive Performance for Clinical and Demographic Features.

The performance increase observed when incorporating scores like the DOCS strongly suggests that these clinical variables, rather than the EEG score itself, were the primary drivers of classification success in these specific combinations. Results showed that certain clinical symptom scores led to substantial increases in performance, particularly the total DOCS (AUC = 0.97), BAI (AUC = 0.94), and BDI-II (AUC = 0.90). The MIS score showed only a modest improvement (AUC = 0.62). Conversely, adding demographic variables (age, sex, education) resulted in performance metrics below the already poor baseline (AUC range 0.18 – 0.38).

## Discussion

This pilot study investigated the feasibility and efficacy of using CNN-based classification on resting-state EEG data to differentiate individuals with OCD from HC. Our primary finding is that the CNN operating on time-frequency representations of EEG signals achieved robust classification performance, significantly outperforming a traditional machine learning baseline model (SVM) trained on standard spectral power features. These results are generally aligned with previous research indicating better predictive power with neural networks (Salomoni et al., 2009) and the need for nonlinear prediction models for OCD (Zaboski et al., 2024). Furthermore, a second-stage analysis integrating the CNN-derived EEG score with clinical and demographic variables revealed that no clinical or demographic variables enhanced classification accuracy, suggesting that all diagnostically relevant signal was captured by the CNN.

The marked contrast between the performance of the CNN and the SVM baseline warrants careful consideration, especially given the non-independent nature of the epoch-level data. As in many EEG studies, CNNs, with their high flexibility, are at risk of overfitting to subject-specific artifacts (e.g., consistent noise patterns or skull conductivity). Conversely, a more rigid SVM trained on handcrafted features may be less sensitive to such noise. An epoch-level comparison alone is therefore insufficient to claim superior performance in detecting disorder-specific neural patterns.

Our primary, subject-level evaluation addressed this challenge by providing a fair comparison of each model’s ability to generalize to unseen individuals. The results of this stringent test were unambiguous: the CNN achieved 85.0% accuracy at the subject level, while the SVM performed no better than chance (45.0% accuracy). This finding strongly suggests that the CNN’s superior performance was not simply an artifact of modeling subject-specific noise. Instead, it indicates a fundamental difference in the utility of the input features: the traditional, averaged spectral band-power features used by the SVM contained little to no generalizable diagnostic information for OCD in this dataset. In contrast, the hierarchical spatio-temporal features automatically learned by the CNN from the complete time-frequency representations contained a robust and generalizable signal sufficient to distinguish individuals with high accuracy.

The baseline model using only the subject-level EEG score derived from the CNN LOSO predictions already performed well (AUC 0.86, Accuracy 80.0%). Contrary to expectations, adding specific symptom scores (DOCS, MIS) or general anxiety/depression scores (BAI, BDI-II), participant age, or sex did not improve classification performance beyond this EEG baseline. This could suggest that the variance captured by these clinical scales was either largely represented within the complex EEG features learned by the CNN, or that these scales did not provide sufficient independent discriminative information at the subject level. Notably, descriptive statistics showed high variability among scores (e.g., DOCS), as well as scores that were similar between groups (BAI). This may have limited their utility for multimodal classification in this cohort due to added noise. While heterogeneity in clinical samples is expected, it underscores the need for replicating our results in larger datasets.

The baseline multimodal performance comparison (CNN vs. SVG) demonstrated the superior discriminative capability of the CNN-derived EEG features over the traditional SVM/band power approach. In sharp contrast, the SVM required the addition of potent clinical symptom scores (DOCS, BAI, BDI-II) to achieve high classification metrics. This improvement largely compensated for the underlying weakness of the SVM’s baseline EEG score, indicating the clinical scores themselves, not the SVM’s limited EEG features, were responsible for the higher performance. Therefore, while the SVM + DOCS combination yielded the highest numerical AUC (0.97) in our analyses, this result primarily reflects the discriminative power of the DOCS score in this sample, not the resting state EEG signals. Conversely, the fact that adding the DOCS score did not improve the already strong CNN baseline (AUC = 0.86) suggests a redundancy between the two features. This implies that the complex, spatio-temporal patterns learned by the CNN from the EEG data already contained the diagnostically relevant information that is captured by the clinical DOCS score—information that was entirely missed by the simpler, band-power-based SVM.

### Strengths and Limitations

Strengths of this pilot investigation include the application of a modern deep learning architecture (CNN) to clinically accessible EEG data, the rigorous LOSO cross-validation approach controlling for individual subject effects, and a direct comparison with a traditional SVM baseline using identical cross-validation. Moreover, we systematically investigated multimodal integration. The preprocessing pipeline also followed established best practices using MNE-Python.

Despite these strengths, we must also acknowledge several limitations. The most significant limitation is the small sample (*n* = 10 per group). While LOSO cross-validation maximizes the use of available data for training in each fold (Mennes et al., 2010; Ren et al., 2024), the overall model generalizability is limited, and the results require replication in larger datasets. The high variability observed in per-fold accuracy (81.29% ± 32.17%) also underscores the influence of individual subject heterogeneity due to sample size. Relatedly, given the pilot nature of this study and the small sample size, which limits the stability and interpretability of confidence intervals for AUC comparisons in the multimodal analysis, we focused on the observed point estimates. These exploratory findings require validation in larger cohorts where more robust statistical inferences, including confidence intervals, can be drawn. Secondly, this study focused solely on resting-state EEG; task-based paradigms might elicit different or more pronounced group differences. Thirdly, the analyses were cross-sectional, using only baseline data, precluding insights into treatment effects or longitudinal changes. Although the CNN performed well, the neural features driving the classification remain relatively opaque—a common “black box” problem with deep learning models.

Additionally, our selection of hyperparameters was based on common practices in the field rather than a systematic optimization search (e.g., grid search). While our chosen parameters yielded strong results, it is possible that further tuning could enhance performance. Similarly, our use of a standard LOSO cross-validation, while rigorous, uses the test set for early stopping. Larger-scale studies should implement procedures such as nested cross-validation frameworks to fully separate hyperparameter tuning from the final model evaluation, though a systematic search and a nested validation approach were considered beyond the scope of this pilot investigation.

### Future Directions

Replication in larger datasets with more diverse demographic and clinical characteristics is paramount to confirm the robustness and generalizability of the CNN’s performance. Applying similar deep learning models to task-based EEG data (e.g., error monitoring, symptom provocation tasks) in OCD could reveal complementary diagnostic information. Utilizing advanced interpretability techniques (e.g., saliency mapping, layer-wise relevance propagation) could help elucidate the specific time-frequency patterns the CNN identified as discriminative (Cui et al., 2023; Lopes et al., 2023). Exploring different deep learning architectures, potentially incorporating temporal dependencies more explicitly, leveraging graph neural networks to model electrode relationships, or using transformer models could yield further improvements (Cisotto et al., 2020; Klepl et al., 2024; Kuruvila et al., 2021; Yao et al., 2024). Longitudinal studies are needed to assess whether these EEG-based models can predict treatment response or track clinical changes over time. Finally, studies are needed to further explore the relationship between the signal captured by EEG and symptom severity.

## Conclusion

This pilot study demonstrates the feasibility of using CNNs applied to resting-state EEG time-frequency data to differentiate individuals with OCD from healthy controls with high accuracy, outperforming a traditional SVM approach based on spectral power. The analysis further suggested that the EEG data alone contained substantial discriminative information, with no other clinical or demographic variables improving model performance—implying that the CNN may be learning the neurophysiological signature of clinical symptom severity itself. Despite the limitations imposed by the small sample size, these findings highlight the potential of deep learning techniques for developing powerful predictive models on complex data types. Further research and validation in larger, more diverse samples are crucial next steps towards translating these advanced computational approaches into tools for research and clinical practice.

## Data Availability

All data produced in the present study are available upon reasonable request to the authors following completion of R01MH116038.

## Supplement 1

This supplement provides the detailed results of the epoch-level classification for the CNN, which formed the basis for the primary subject-level analysis presented in the main manuscript. When predictions were aggregated across all 2,277 test epochs from the 20 held-out subjects, the CNN’s overall classification accuracy reached 81.95%, and the ROC AUC was 0.86. Detailed performance metrics, including precision, recall, and F1-score, are presented in Table S1.

**Table S1:** Overall Classification Performance of the CNN at the Epoch Level.

| **Class** | **Precision** | **Recall** | **F1-Score** | **Support** |
| --- | --- | --- | --- | --- |
| HC | 0.84 | 0.79 | 0.81 | 1148 |
| OCD | 0.80 | 0.85 | 0.82 | 1129 |
| **Accuracy** | - | - | 0.82 | 2277 |
| **Macro Avg** | 0.82 | 0.82 | 0.82 | 2277 |
| **Wtd Avg** | 0.82 | 0.82 | 0.82 | 2277 |

*Notes:* F1-Score: A single metric that balances precision and recall; support: Number of epochs belonging to that true class in the aggregated test sets; accuracy: Proportion of correctly classified epochs; macro avg: The unweighted average of precision, recall, F1 across both classes; weighted avg: The average of the metric across both classes, weighted by the support (number of true instances) for each class.

**Figure S1:**
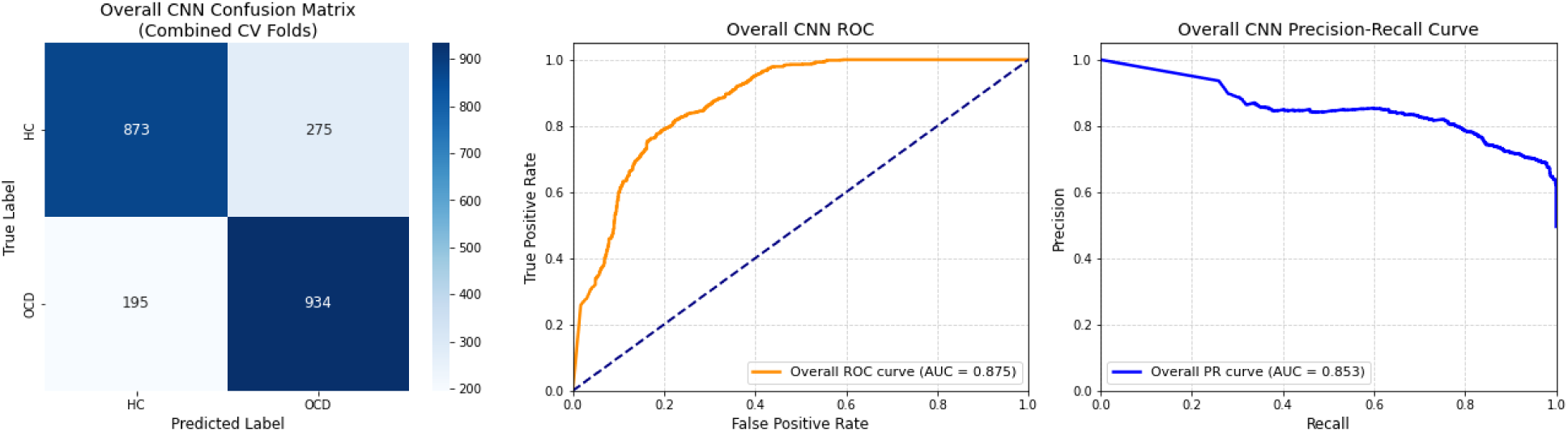
CNN Confusion Matrix, Receiver Operator Curve (ROC), and Precision-Recall Curve.

The model’s overall epoch-level performance characteristics are illustrated in Figure S1. The confusion matrix details the specific classification outcomes across all test epochs: 873 True Negatives (HC epochs correctly identified), 934 True Positives (OCD epochs correctly identified), 195 False Negatives, and 275 False Positives. The ROC and Precision-Recall curves further illustrate the model’s performance on this epoch-level task.

**Figure S2:**
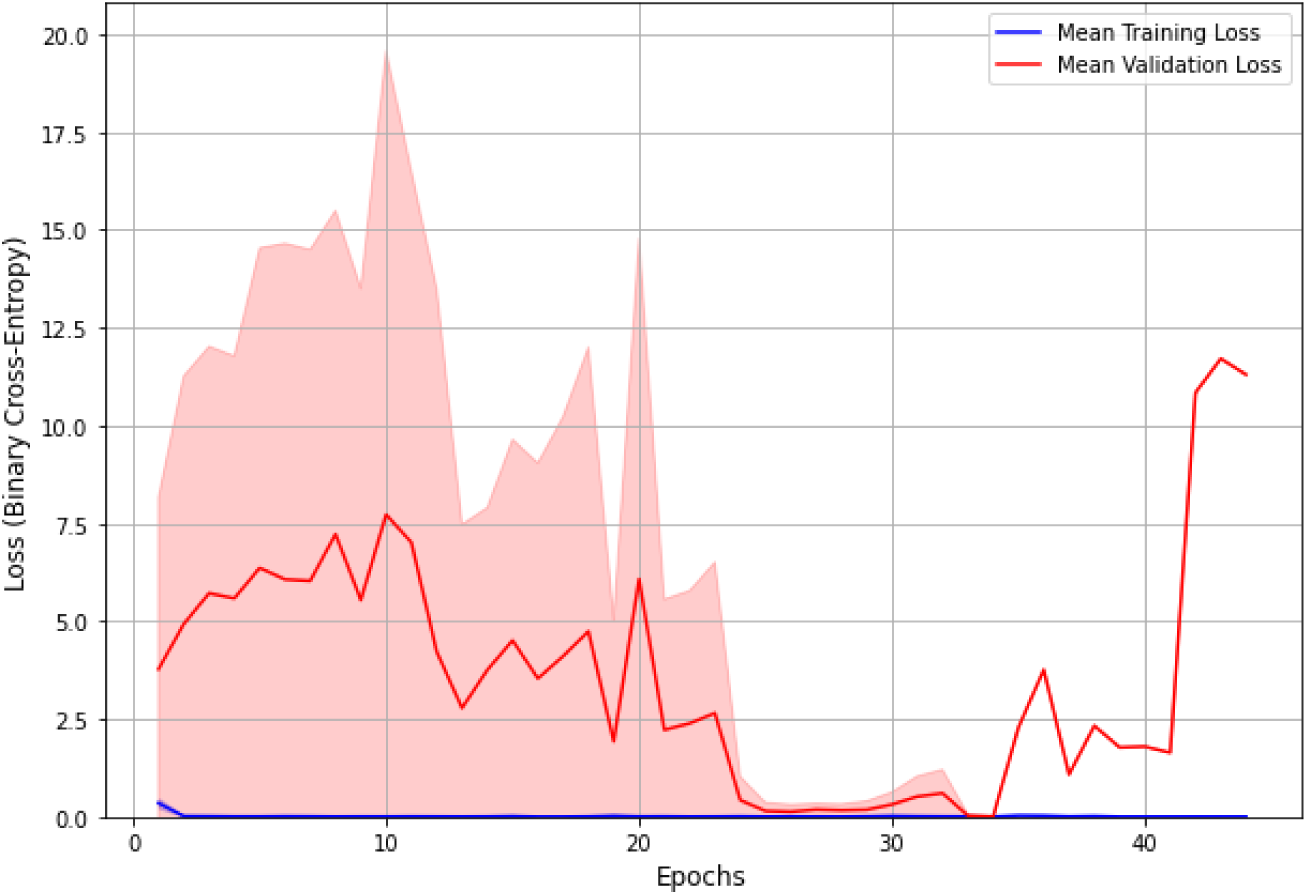
Average Training and Validation Loss.

Figure S2 shows the mean loss (solid lines) and standard deviation (shaded areas) across all 20 folds of the leave-one-subject-out cross-validation. The training loss (blue) rapidly converges, while the validation loss (red) also decreases, indicating model generalization. The high variance in the validation loss reflects the significant inter-subject heterogeneity in the data and the need for the subject-level analysis.

## Supplement 2

This supplement provides the results of the epoch-level classification for the SVM, which formed the basis of comparison with the CNN. For comparison, we constructed an SVM trained on spectral power features (delta: 1 – 4 Hz, theta: 4 – 8 Hz, alpha: 8 – 13 Hz, beta: 13 – 30 Hz, gamma: 30 – 45 Hz). When aggregated across all 2,277 test epochs, the SVM’s epoch-level accuracy was 49.01%, and the ROC AUC was 0.449. The full epoch-level model metrics are presented in Table 3, and the corresponding confusion matrix is shown in Figure S3.

**Table S2.** Overall Classification Performance of the SVM at the Epoch Level.

| **Class** | **Precision** | **Recall** | **F1-Score** | **Support** |
| --- | --- | --- | --- | --- |
| HC (0) | 0.49 | 0.52 | 0.51 | 1148 |
| OCD (1) | 0.48 | 0.46 | 0.47 | 1129 |
| **Accuracy** | - | - | 0.49 | 2277 |
| **Macro Avg** | 0.49 | 0.49 | 0.49 | 2277 |
| **Wtd Avg** | 0.49 | 0.49 | 0.49 | 2277 |

*Notes:* F1-Score: A single metric that balances precision and recall; support: Number of epochs belonging to that true class in the aggregated test sets; accuracy: Proportion of correctly classified epochs; macro avg: The unweighted

**Figure S3:**
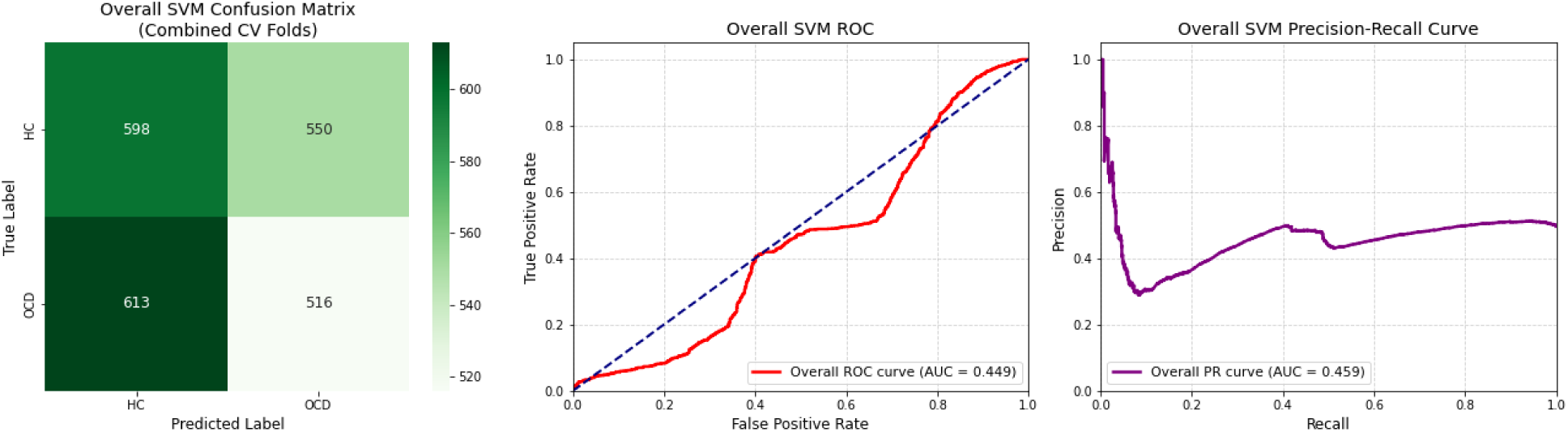
SVM Epoch-Level Confusion Matrix, ROC, and Precision-Recall Curve.

